# A Randomized Pilot Trial of Medically Tailored Meals and Lifestyle Support for Gestational Diabetes: Feasibility, Acceptability, and Implementation Challenges

**DOI:** 10.64898/2026.04.16.26351041

**Authors:** Andrea D. Shields, Molly E. Waring, Makayla Murphy, Linda S. Pescatello, Ock K. Chun, Helen Wu, Vanessa Sena, Christiana M. Field, Annie Kearns

## Abstract

**Background:** Lifestyle interventions incorporating medically-tailored meal delivery may support rapid behavior change among pregnant individuals with gestational diabetes (GDM).

**Purpose:** To examine the feasibility and acceptability of a multicomponent lifestyle intervention for pregnant individuals with GDM. Primary outcomes included recruitment, retention, intervention receipt, and acceptability.

**Methods:** We conducted a pilot randomized feasibility trial among pregnant individuals with GDM recruited from maternal fetal medicine clinics in the Hartford, Connecticut area. Participants were randomized to usual GDM care or the Meals4Moms intervention plus usual care. The intervention included medically-tailored meal delivery, personalized physical activity support, and multimodal education with digital tools. Participants completed a survey and three 24-hour dietary recalls at baseline and post-intervention. Meals4Moms participants also completed a semi-structured interview at follow-up. Intervention receipt was tracked by study staff.

**Results:** Of 30 individuals approached, we screened 80% (n=24), of whom 75% (n=18/24) were eligible; we randomized 8 participants. Seventy-five percent (n=6/8) completed at least one component of the follow-up assessment (100%, n=4/4 Meals4Moms, 50%, n=2/4 Usual Care). One participant spent ≥80% of her total food budget (n=1/4, 25%), and no participants completed ≥80% of prescribed exercise sessions (range: 0-50%). All (n=4) Meals4Moms participants reported they would be very likely to participate in the program if they had GDM again, and 100% (n=4) would be very likely to recommend the program to a friend with GDM.

**Conclusions:** While the Meals4Moms intervention was highly acceptable to participants, procedural refinements are needed prior to conducting a full-scale efficacy trial.

## Introduction

Gestational diabetes mellitus (GDM) affects approximately 2–10% of pregnancies in the United States and is associated with a range of maternal and neonatal complications [1]. Compared with pregnancies without GDM, affected individuals have higher risk of adverse outcomes, including antepartum hospitalization, cesarean delivery (50% vs 35%), and longer hospital stays [2–5]. Infants born to persons with GDM are at increased risk for metabolic and hematologic disorders, respiratory distress, cardiac complications, perinatal asphyxia, birth trauma, and neonatal intensive care unit (NICU) admission [6,7]. In addition, nearly half of individuals diagnosed with GDM will develop Type 2 diabetes mellitus later in life, significantly increasing their lifetime risk of cardiovascular disease (CVD) [1].

Given these long-term health implications, a GDM diagnosis represents a critical opportunity for early intervention—not only to improve pregnancy outcomes but also to promote lasting lifestyle changes that benefit both the pregnant individual and their offspring. However, effective management of GDM requires multifaceted, patient-centered approaches that provide that combine medical care, education, and behavioral support during and after pregnancy. Standard GDM management includes adherence to American Diabetes Association (ADA)-recommended dietary recommendations, regular glucose monitoring, physical activity, and prenatal care compliance [2–5]. Unlike individuals with prediabetes or Type 2 diabetes, individuals with GDM must adopt these changes rapidly - typically within 8–10 weeks before delivery - requiring immediate access to nutrient-dense foods and practical education on meal planning, portion control, and healthy snacking. While food prescription programs have improved glycemic outcomes in populations with Type 2 Diabetes [8], similar interventions tailored to GDM remain unexplored. Exercise interventions have also demonstrated benefits for glycemic control in GDM, but most rely on in-person supervision, which limits scalability [9–12]. To improve accessibility and sustainability, interventions should emphasize self-efficacy, remote support, and behavioral strategies such as goal setting, progress tracking, and personalized feedback [13].

To address these gaps, we developed the Meals4Moms program, an innovative lifestyle intervention that integrates: 1) immediate access to GDM-compliant, medically-tailored meals (MTM-GDM) along with multimedia nutrition education (e.g., recipes and cooking demonstrations led by community-based culinary experts); 2) remote, asynchronous exercise support using wearable devices and personalized feedback from trained staff; and 3) comprehensive digital education resources, including podcasts, FAQs, and expert-authored blog content to support ongoing engagement [14]. By combining nutrition, physical activity, and digital education, Meals4Moms is designed as a scalable, patient-centered intervention to improve both short-term pregnancy outcomes and long-term cardiometabolic health. The objective of this study was to conduct a pilot randomized feasibility trial to evaluate the feasibility and acceptability of the Meals4Moms intervention compared with usual care among pregnant individuals with GDM.

## Methods

We conducted a pilot randomized feasibility trial comparing the Meals4Moms lifestyle intervention plus usual care versus usual care for pregnant individuals with GDM. Primary feasibility outcomes were recruitment, retention, intervention receipt, and acceptability. The study was approved by the UConn Health Institutional Review Board (IRB# 23-190SSF-2) and pre-registered at https://clinicaltrials.gov (NCT06227247).

Eligible participants were aged 18 to 49 years, pregnant with a singleton gestation, diagnosed with GDM between 24+0 and 31+6 weeks gestation, and within 4 weeks of GDM diagnosis at enrollment. Participants received care at the UConn Health Maternal Fetal Medicine Clinic, St. Francis Hospital Women’s Health clinic, or Hartford Hospital, and planned to deliver at these institutions. Addition requirements included English proficiency, internet access, medical clearance for physical activity, and residence within the meal delivery area. Individuals were excluded if they had pregestational diabetes mellitus, were diagnosed with GDM outside the eligibility window, were scheduled for a medically-indicated preterm birth (i.e., placenta accreta, prior classical incision), had conditions limiting participation (e.g., inability to tolerate solid food), were enrolled in another intervention study, or had dietary restrictions that could not be accommodated. While individuals of all gender identities were eligible to participate, all enrolled participants identified as female, and thus we refer to participants as “women”.

Research staff screened interested individuals by phone, obtained permission for chart review, and conducted informed consent electronically via REDCap. Recruitment occurred from February 2024 through January 2025.

Participants were randomized 1:1 to the Meals4Moms versus Usual Care conditions using permuted blocks (size 4 and 6) stratified by site (UCHC vs SFH vs HH) and presence of children in the household (0 children vs 1+ children) to balance potential differences in clinical care and experiences changing diet due to family configuration, respectively. Randomization was conducted through REDCap by study staff using a randomization table generated by the study statistician (MEW) [15]. Staff informed participants of their treatment assignment via phone.

Usual GDM care (Usual Care) was provided by the participant’s prenatal care provider. Usual GDM care generally consists of one visit with a registered diabetes educator to review the GDM diet, education on glucometer use and monitoring of fasting and postprandial blood glucose levels by the maternal-fetal medicine team, and encouragement/guidance of incorporating or increasing physical activity by the patient’s obstetric and maternal-fetal medicine clinicians [16]. Depending on pregestational weight, weight management may also be incorporated into treatment [16]. Further, pharmacotherapy (i.e., insulin) is an integral component of treatment if GDM is not manageable through lifestyle modification alone. In terms of a GDM diet, nutrition counseling typically endorses a balance of macronutrients [16]. Medical nutrition therapy initiated through a referral to a registered dietitian-nutritionist (RD/RDN) supports the establishment of a food plan, weight goals, and an appropriate insulin-to-carbohydrate ratio as applicable [16]. Individuals are typically encouraged to engage in both aerobic activity and strength training and increase activity up to 150 minutes of moderate-to-vigorous intensity physical activity per week [17,18].

Participants randomized to the Meals4Moms condition received usual GDM care plus the Meals4Moms, a multicomponent, community-based, healthy-living lifestyle intervention including continued GDM education, physical activity level monitoring, and delivery of MTMs-GDM for the management of gestational diabetes in pregnant people. The program was centered around providing a subsidized food prescription program designed to promote the consumption of nutritious GDM-specific meals. Every week, My Local Chefs, a local Connecticut-based small business, packaged and delivered prepared MTMs-GDM, along with nutritionist-approved GDM-tailored recipes that guided participants to make healthy meals at home. Meals were locally-sourced, culturally diverse, and prepared with nutrient-rich foods. Participants randomized to the Meals4Moms condition received an individualized link to the My Local Chefs website, where they could select meals each week. Participants were provided a budget of $266 per week (equivalent to 7 breakfasts, 7 lunches, and 7 dinners), but were allowed to customize their orders to their dietary preferences and family food systems. My Local Chefs also offered bags of fresh produce (small and large bags) that participants could include in their orders. Prepared meals were delivered to the participant’s home from their first order post-randomization until 1 week after postpartum (on average, 8 to 11 weeks). Prepared meals included the recipes, tips for meal preparation, and meal-specific nutrition information. Participants submitted meal orders on Mondays, except for weeks with a Monday federal holiday in which participants submitted orders on the preceding Friday or following Tuesday (e.g., Friday before Memorial Day, Tuesday after Labor Day).

Participants initially met virtually or over the phone with the study’s exercise trainer to discuss their baseline exercise level and health status to inform the development of personalized fitness goals during pregnancy. Led by study kinesiologist (LSP), the team created two exercise videos specifically for the Meals4Moms intervention. The exercise trainer made recommendations to which videos were appropriate for each participant based on participants’ baseline fitness levels. Participation in the exercise videos was unsupervised. Participants were instructed to use Move Your Way® from the Office of Disease Prevention and Health Promotion, a free online customizable weekly physical activity planner to set exercise goals and track progress [17]. Participants were provided with an activity tracker (Fitbit Inspire 2) to monitor their daily physical activity and a Wi-Fi scale (Fitbit Aria Air) to monitor gestational weight gain. Participants were asked to wear the Fitbit as often as possible and to self-weigh daily. Every two weeks, the exercise trainer met remotely with each participant to review their progress and set new goals. Before these sessions, the trainer accessed a dashboard on the Fitabase digital platform (Fitabase, San Diego, CA) to review each participant’s activity data. Via the intervention website, participants had access to multi-modal educational materials on exercise, nutrition and blood glucose management, including videos, podcasts created by the study PI (ADS), blog posts, and other educational materials. My Local Chefs chefs created cooking demonstration videos corresponding to MTMs-GDM offered during the study. In addition to the two exercise videos created by the Meals4Moms study team, participants could access a library of links to exercise and yoga videos available for free online. This library was developed as part of our previous work with pregnant populations [19], and videos were re-reviewed for safety for individuals with GDM by the study PI (ADS) and kinesiologist (LSP).

At baseline, participants completed an online survey via REDCap [20] and three 24-hour dietary recalls using the National Cancer Institute’s (NCI) automated self-administered 24-hour dietary recall (ASA24™; version ASA24-2022) [21]. Research staff obtained medical clearance from participants’ prenatal care teams. Participants in both treatment conditions completed a follow-up survey via REDCap between 37 weeks’ gestation and 2 weeks post-delivery. The follow-up survey included many of the measures assessed at baseline. Participants also rated how acceptable they found their experience with the study [22]. Participants randomized to the Meals4Moms condition were also asked about their satisfaction with meals and acceptability of the program. Participants in both treatment conditions completed three 24-hour dietary recalls via the ASA24 online system (2 weekdays and 1 weekend day) using a similar procedure as at baseline. Participants in the Meals4Moms condition completed a semi-structured interview via video conferencing software (WebEx).

Participants in both study conditions were compensated for their time to complete research assessments (up to $100 in gift cards). Participants received a $20 gift card for completion of the assessments at each timepoint (baseline and follow-up) and a $10 gift card for each completed diet recall at baseline and follow-up assessment timepoints (up to $60 if completed all 6 recalls). To receive the gift card at follow-up, participants in the Meals4Moms condition had to complete both the online survey and interview. Meals4Moms participants were allowed to keep the activity tracker and Wi-Fi scale.

Clinical outcomes were abstracted from participants’ electronic health record into REDCap by study staff. Clinical data included maternal characteristics (e.g., age, gravity, parity), preexisting medical conditions and medication use, detailed GDM management outcomes (e.g., use of insulin), and pregnancy and neonatal outcomes. Study staff downloaded activity and weight data from Fitabase [23]. Data was routinely synced and stored from the time participants received their devices to the date they completed follow-up procedures. My Local Chefs staff provided data on weekly meal orders.

The primary outcomes of this feasibility pilot trial were recruitment, retention, intervention receipt, and intervention acceptability. Research staff tracked participants through study procedures, including recording reasons for ineligibility and non-participation. We calculated recruitment from the number of individuals approached, eligible, and randomized, and calculated recruitment pace. Our a priori feasibility benchmark for recruitment was ≥75% of eligible participants enrolled. We measured retention as the proportion of participants who completed any aspect of the follow-up assessment (benchmark: ≥80%). As our goal was to complete follow-up within 2 weeks post-delivery, we conducted a sensitivity analysis in which we assessed retention as the completion of any aspect of the follow-up assessment within 2 weeks post-delivery. We assessed intervention receipt as a) the percent of Meals4Moms intervention participants who spent at least 80% of their weekly $266 food budget and b) the percent of participants who completed at least 80% of sessions with the fitness trainer that they were eligible to complete. Participants assigned to the Meals4Moms study condition were referred to My Local Chefs immediately following randomization to the Meals4Moms condition. Following the intervention, My Local Chefs provided order data to the research team (i.e., order date, items ordered, cost per items). As meal orders were due on Mondays, we calculated the eligible weekly orders as the number of Mondays between randomization and delivery. We estimated a total eligible food budget as $266 weekly food budget multiplied by the number of meal orders the participants were eligible to submit, and calculated the proportion of participants who spent at least 80% of their total food budget. We also calculated the proportion of orders using at least 80% of the weekly food budget. Research staff recorded the number of exercise sessions each participant completed. We calculated the number of exercise sessions each participant was eligible to complete by assuming that the first session would occur within 7 days of randomization and then every 2 weeks (14 days) until delivery. From this, we calculated the proportion of participants who completed at least 80% of their eligible exercise sessions.

Intervention acceptability included two dimensions: a) the percentage of participants who reported they would be likely or very likely to participate again if they had recurrent diagnosis of GDM and b) the percent of participants who reported they would be likely or very likely to recommend the Meals4Moms intervention to a friend with GDM. A priori benchmarks for these metrics were ≥80% responding “likely” or “very likely” to these acceptability questions. These acceptability questions [19,22] were included in the follow-up survey (response options: very unlikely, unlikely, neutral, likely, and very likely). Secondary acceptability questions included likelihood of recommending the meal delivery and the physical activity support components of the intervention to a friend with GDM, frequency of use/engagement with different intervention components (e.g., podcasts, recipes; response options: not at all, once or twice, more than twice but not every week, at least once a week), and among those reporting use of intervention components, how helpful and then relevant this component was to their life (response options: not at all, a little bit, somewhat, quite, very helpful/relevant). In post-intervention interviews, participants were asked what they found most and least helpful about the Meals4Moms intervention, suggestions for improvement, and then their experiences with and thoughts about the different components of the intervention (e.g., meal delivery, intervention website, exercise sessions with trainer).

At eligibility screening, participants reported their age in years and gestational age in weeks. On the baseline survey, participants reported demographic and household characteristics. Participants were asked to select which race(s) best describes themselves and reported whether they consider themselves as Hispanic or Latina. Participants rated how difficult it has been for their household to pay for usual household expenses (e.g., food, rent or mortgage, car payments) [24] and whether anyone in their household received benefits from ‘Temporary Assistance for Needy Families (TANF)’, or ‘SNAP or Food Stamps (including pandemic-EBT or P-EBT)’, ‘Women, Infants, and Children (WIC) program’ or ‘Other food assistance program (Commodity Supplemental Food program, Meals on Wheels, or other)’ in the past 12 months. Participants completed the 18-item USDA food security screener [25]. Health literacy was assessed using the Newest Vital Sign: participants were asked to read a nutrition label and answer questions [26]. We summed the total score and categorized participant’s health literacy as: impaired health literacy (0-3 items correct) or adequate health literacy (4-6 items correct) [26]. Participants reported whether anyone in their household followed a special diet or had food allergies.

Participants self-reported their height and pre-pregnancy weight. We calculated pre-pregnancy BMI and categorized as underweight (BMI <18.5 kg/m^2^), normal weight (18.5 kg/m^2^ ≥ BMI < 25 kg/m^2^), overweight (25 kg/m^2^ >BMI < 30 kg/m^2^), and obesity (30 kg/m^2^ ≥ BMI) [27]. Participants reported the frequency and duration in a typical week they engaged in vigorous-intensity exercise, moderate-intensity exercise, or walking [28].

Participants reported if they ever had meal kits (e.g., Blue Apron or HelloFresh) delivered and if so, whether they currently have meal kits delivered. Participants reported whether they own a smartphone (e.g., iPhone or Android phone), whether they have ever used Wi-Fi scale (e.g., Fitbit Aria or the Withings scale), and if they have ever used a fitness tracker (e.g., Fitbit) to track their steps or exercise. Participants were asked whether they have subscribed to any podcasts and if they have subscribed, they were asked how often they listened to podcasts in the past 7 days.

As elevated depressive symptoms are common during pregnancies complicated by GDM [29], we included a measure of depression in our baseline and follow-up surveys, the 10-item Edinburgh Postnatal Depression Scale (EPDS) [30]. Study staff reviewed EPDS scores within 24 hours of survey completion. Research staff reported scores ≥12 to the PI (ADS). The PI proceeded to contact the participant to discuss score and recommend mental health care prior to notifying their prenatal provider. These steps were documented in the participant’s EHR and study tracking database. If participants scored positive on the self-harm question, they were considered at acute risk of injury or harm. In such cases, the project manager would alert the PI immediately, and the participant would be assessed for immediate referral for psychiatric evaluation. The participant’s prenatal provider would be made aware of the situation.

### Statistical Analyses

We had aimed to recruit 40 pregnant individuals with GDM. This target sample size was based on numbers needed to determine feasibility (particularly of recruitment pace) and informed by availability of funding for this feasibility pilot study.

We used Research Electronic Data Capture (REDCap) [20] for participant tracking and administering participant surveys. We conducted data management and descriptive analyses using SAS 9.4 (SAS Institute, Inc., Cary, NC). We summarized participant characteristics and feasibility and acceptability outcomes, overall and by treatment condition. We reported recruitment and retention using a CONSORT diagram specific to feasibility pilot trials [31]. We exported transcripts auto-generated by WebEx to Excel. One member of the research team listened to the recording and corrected the transcript, and a second team member listened to the recording to confirm the corrections. We conducted a rapid qualitative analysis [32] of participant feedback to inform continued development of the Meals4Moms program [14].

## Results

### Recruitment

We approached 30 pregnant individuals with GDM, of whom 24 completed eligibility screening; 18 were eligible (Figure 1). We randomized 8 participants (n=4 Meals4Moms, n=4 Usual Care; Figure 1). Baseline characteristics are shown in Table 1 and Supplemental Table 1. All (n=8) were married or living with a partner; one participant randomized to the Meals4Moms condition lived with an additional adult. One participant (Meals4Moms) received WIC benefits; none received TANF or SNAP. All (n=8) had adequate health literacy. A quarter (n=2; n=1 per condition) reported a household member following a special diet and 25% (n=2; n=1 per condition) reported food allergies. Half (n=4, 50%) had ever previously used a meal kit delivery service (n=3, 75% Meals4Moms, n=1, 25% Usual Care). One participant in the Usual Care condition reported using a meal kit delivery service in the current pregnancy.

**Figure 1:**
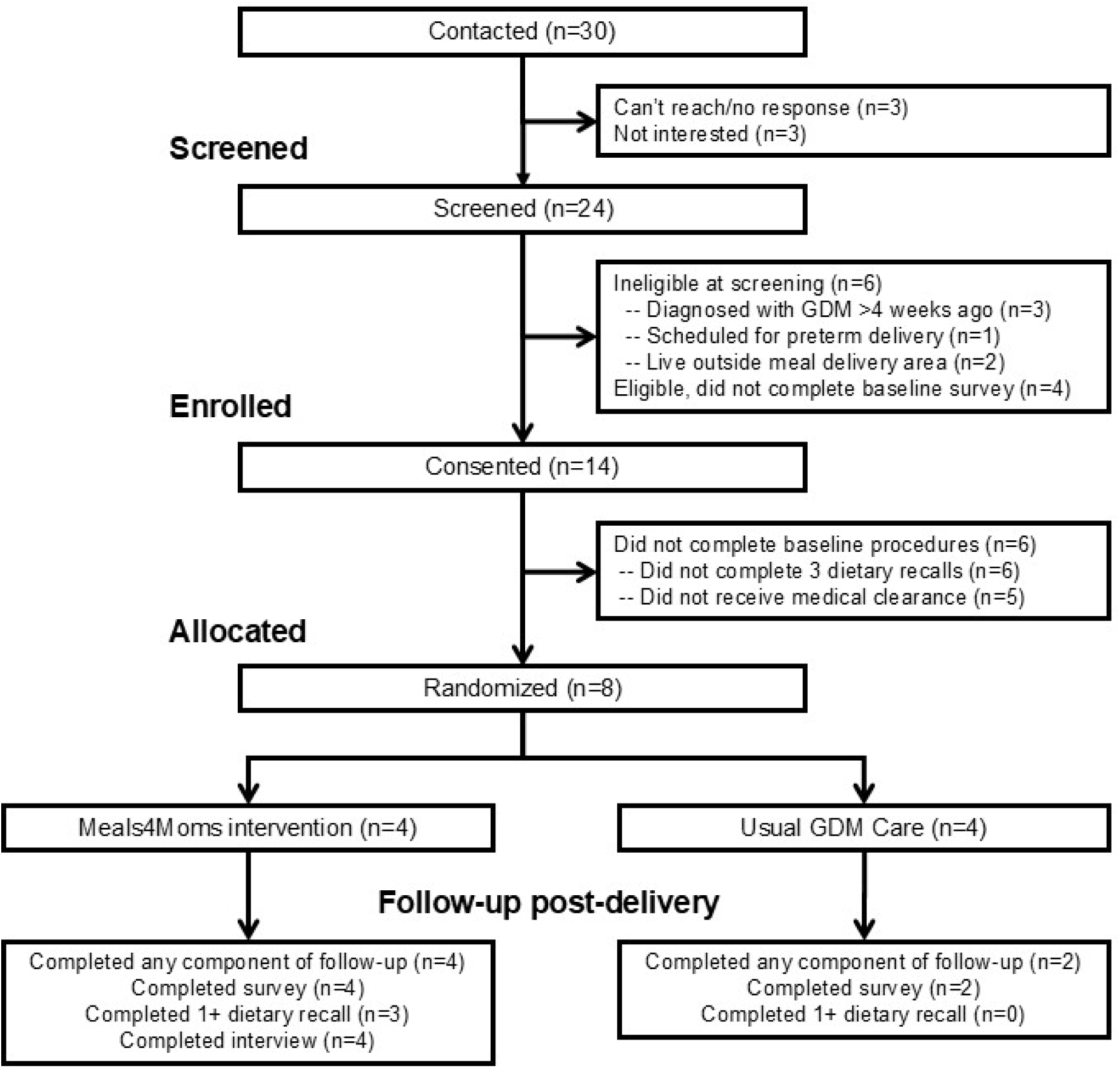
CONSORT diagram.

**Table 1:** Characteristics of pregnant women with gestational diabetes (GDM), overall and by study condition, n (%) or M±SD.

|  | Overall<br>(n=8) | Meals4Moms<br>(n=4) | Usual care<br>(n=4) |
| --- | --- | --- | --- |
| Age (years) | 33.0±4.2 | 35.5±3.9 | 30.5±3.0 |
| Weeks pregnant at eligibility screening | 30.1±1.1 | 29.3±0.5 | 31.0±0.8 |
| Race and ethnicity |  |  |  |
| Non-Hispanic white | 7 (83) | 4 (100) | 3 (75) |
| Hispanic/Latina (white race) | 1 (13) | - | 1 (25) |
| Education |  |  |  |
| High school/GED | 1 (13) | - | 1 (25) |
| 1-3 years of college or Associate's degree | 1 (13) | - | 1 (25) |
| Bachelor's degree | 1 (13) | 1 (25) | - |
| Graduate degree | 5 (63) | 3 (75) | 2 (50) |
| Employment status |  |  |  |
| Working full-time | 6 (75) | 3 (75) | 3 (75) |
| Working part-time | 1 (13) | - | 1 (25) |
| Stay-at-home mom | 1 (13) | 1 (25) | - |
| Number of children in household |  |  |  |
| 0 children | 5 (63) | 2 (5) | 3 (75) |
| 1 child | 2 (25) | 1 (25) | 1 (25) |
| 2 children | 1 (13) | 1 (25) | - |
| Difficulty paying for basic expenses |  |  |  |
| Not at all difficult | 4 (50) | 3 (75) | 1 (25) |
| A little difficult | 2 (25) | - | 2 (50) |
| Somewhat difficult | 2 (25) | 1 (25) | 1 (25) |
| Very difficult | - | - | - |
| Food security |  |  |  |
| High food security | 6 (75) | 3 (75) | 3 (75) |
| Marginal food security | - | - | - |
| Low food security | 1 (13) | - | 1 (25) |
| Very low food security | 1 (13) | 1 (25) | - |
| Any vigorous physical activity in past 7 days at baseline* | 1 (14) | - | 1 (25) |
| Any moderate physical activity in past 7 days at baseline | 5 (63) | 3 (75) | 2 (50) |
| Household member follows special diet |  |  |  |
| Self (not including GDM diet) | 2 (25) | 1 (25) | 1 (25) |
| Another adult | 1 (13) | 1 (25) | - |
| Child(ren) (n=3) | 1 (33) | 1 (50) | - |
| Household member has food allergy |  |  |  |
| Self | - | - | - |
| Another adult | - | - | - |
| Child(ren) (n=3) | 2 (67) | 1 (50) | 1 (100) |

\* Missing data: n=1 for any vigorous physical activity in past 7 days at baseline

Recruitment occurred from February 2024 through January 2025. Over this 11-month period, our monthly recruitment pace was 2.7 approached, 2.2 screened, 1.6 eligible, and 0.7 randomized. The trial was stopped early due to insufficient recruitment to support a full-scale efficacy trial.

Most randomized participant identified as non-Hispanic white (n=7/8, 88%) compared to 50% of eligible individuals who consented but did not complete baseline procedures (n=3/6, 50%; n=3 non-Hispanic white, n=2 non-Hispanic Asian, n=1 Hispanic/Black). All randomized participants had adequate health literacy (n=8/8, 100%), whereas only half of non-randomized eligible individuals did (n=2/4, 50%). Other characteristic, including partner status, receipt of financial/nutrition assistance, and food security, appeared similar between groups.

### Retention

Seventy-five percent of participants (n=6/8) completed at least one component of the follow-up assessment (n=4, 100% Meals4Moms; n=2, 50% Usual Care; Figure 1). No Usual Care participants completed dietary recalls at follow-up (n=0, 0%). In the Meals4Moms condition, one participant completed all 3 recalls, one completed 2 recalls, one completed a single recall, and one complete none. Only one participant completed all components of follow-up (n=1, 25% in Meals4Moms, n=0, 0% in Usual Care). Although follow-up was intended to occur within 2 weeks postpartum, only 38% (n=3) of participants completed at least one component any follow-up within this timeframe (n=3, 75% Meals4Moms; n=0, 0% Usual Care). One Usual Care participant completed follow-up when 3 months postpartum after returning to work; she had provided her work email for study contacts.

### Intervention receipt

Participants submitted 2-6 meal orders (50-100% of eligible orders) and spent $526.57-$1325.81 corresponding to 49%-97% of their total eligible food budget. Only one participant spent at least 80% of her total eligible food budget (n=1, 25%). However, 94% of orders (n=15/16) were for more than 80% of the weekly budget of $266. The only order that did not near participants’ weekly budget occurred when a participant missed the order deadline, but the My Local Chefs team was able to provide her with a family-sized produce bag ($60). All participants spent at least 80% of their food budgets based on submitted orders (83% to 99% of total food budgets; n=4, 100% of participants). Missed orders most commonly occurred during the first week post-randomization or the final week before delivery. One participant experienced delays due to technical issues and vendor relocation. In one care, a participant submitted her final meal order early on a Tuesday for the following week, and underwent induction of labor later that week, resulting in a post-partum meal delivery.

Participants were eligible to complete 2-4 exercise sessions. Two participants completed one session each, and two completed none. No participants met the threshold of completing ≥80% of eligible (range: 0% to 50%). The first randomized participant was not seen by the trainer due to a communication error. Moreover, an approximately year-long delay in initiating enrollment led to changes in the trainer’s availability. To address this, the study team, including the PI (ADS), met with the trainer to review logistics, clarify the protocol, and align expectations.

### Intervention acceptability

All Meals4Moms participants (100%, n=4) reported that they would be very likely to participate again and to recommend the Meals4Moms program to a friend with GDM. All participants reported that they would be very likely to recommend the meal delivery part of the Meals4Moms program to a friend with GDM. In contrast, only half (n=2, 50%) would be likely or very likely to recommend the physical activity support part of the Meals4Moms program to a friend with GDM (n=1, 25% very unlikely, n=1, 25% neutral, n=1, 25% likely, n=1, 25% very likely). Use of intervention components and ratings of helpfulness and relevance to participants’ lives varied (Supplemental Table 2). In the post-intervention interviews. All participants identified meal delivery as the most helpful component, citing time burden and easier diet management. Least helpful aspects of the intervention, included technical issues with the intervention website and/or meal ordering system, difficult coordinating with the exercise trainer, and limited meal options for some dietary preferences.

### Adverse events

One participant was hospitalized for kidney stones during baseline procedures; this event was unrelated to the study. At baseline, 2 participants reported elevated depression symptoms (n=2 / 14; 14%), one of whom also responded “hardly ever” to the self-harm item. At follow-up, one participant reported elevated depression symptoms (n=1 / 6; 17%), consistent with baseline findings.

## Discussion

Despite high acceptability of the Meals4Moms intervention, several logistical and technical challenges limited feasibility, particularly related to recruitment. We discontinued the trial before reaching our target sample size because the recruitment pace (8 randomized participants over 11 months) was insufficient to support a future efficacy trial without procedural changes. Nonetheless, this pilot provided critical insights to inform refinement of both the intervention and study design.

The primary barrier to feasibility was participant burden during enrollment, particularly dietary recall requirements, rather than lack of interest in the intervention. Completion of 3 dietary recalls at baseline was burdensome and contributed to attrition among 42% of consented participants (n = 6 / 14). This loss appeared more common among participants from racial/ethnic minority groups and those with limited health literacy. Follow-up dietary recall completion was low (13%, n = 1 / 8 completed all 3 recalls). Engagement with the physical activity component was also limited due to structural delivery problems. Delays between eligibility screening and randomization (median 24 days) likely reduced early engagement. Future studies will allocate additional staff resources to support recruitment, enabling more consistent coordination and communication with clinical teams at participating sites. To reduce participant burden, dietary recalls will be offered via telephone. Protocols will also be revised to facilitate earlier engagement with the exercise trainer, with the goal of improving participation before physical limitations later in pregnancy arise [33].

Despite these feasibility challenges, intervention acceptability was high. Participants valued the convenience of medically-tailored meals, predictable portioning, and alignment with clinical guidance, all of which reduced the cognitive burden of dietary management during pregnancy [14]. These findings are consistent with broader literature demonstrating that medically-tailored meal and meal-kit programs improve diet quality, food security, and self-efficacy, and may improve glycemic outcomes among individuals with type 2 diabetes (T2DM) [8]. Evidence from produce prescription and medically-tailored meals programs suggests higher engagement and retention when delivered foods are culturally acceptable and accompanied by nutrition coaching or behavioral supports [34,35]. While formal trials in GDM are limited, early feasibility pilots indicate that structured, lower glycemic load meal kits can improve adherence to dietary recommendations and patient-reported confidence in managing glucose targets [36–38], especially when paired with remote glucose monitoring and app-based feedback [39,40]. Taken together, these data underscore the promise of an integrated model that includes medically-tailored meals/meal kits plus remotely-delivered information and support to improve outcomes in pregnancies complicated by GDM. These findings also suggest differential feasibility across intervention components, with meal delivery highly feasible but exercise support requiring redesign.

This study has important limitations. The sample was predominantly non-Hispanic White (88%), highly educated (75% with a Bachelor’s degree), and included few participants receiving WIC benefits (13%), which does not reflect the broader diversity of birthing people in the United States [27–29]. The homogeneity of this sample limits external validity and may reduce the generalizability of our findings to populations disproportionately affected by GDM, including racially and ethnically diverse and lower-income groups [41]. Future studies should prioritize recruitment strategies that enhance diversity, including partnerships with community-based clinics, WIC programs, and Medicaid populations, and should adapt materials to be culturally and linguistically appropriate.

Beyond individual behavior change, the findings from this feasibility pilot trial have broader implications for how health systems and policy. MTM programs represent a promising “food as medicine” strategy that addresses both clinical care and social drivers of health [42]. Integrating MTMs into existing maternal health programs - such as WIC, Medicaid perinatal care models, and hospital-based diabetes education - may improve equity, sustainability, and cost-effectiveness [43]. For example, WIC already provides nutritional education and food vouchers to low-income pregnant individuals. Partnerships could allow for bundled delivery of clinically appropriate MTMs or meal kits during the high-risk perinatal period. Medicaid programs, many of which now cover nutrition and home-delivered meals as part of value-based or Section 1115 waiver models, offer a promising reimbursement pathway for MTM programs that demonstrate reductions in costly complications such as insulin initiation, hypertensive disorders, or neonatal intensive care use [44]. Similarly, embedding meal support within hospital-based GDM or obstetric nutrition programs could streamline referral and monitoring, integrating meal delivery data into electronic health records for care coordination.

From a policy perspective, demonstrating feasibility and acceptability in pregnancy is a critical step toward expanding coverage of nutrition interventions in maternal health. Future research should evaluate clinical outcomes, cost-effectiveness, and healthcare utilization to support sustainable implementation. By aligning clinical innovation with social policy levers, interventions like Meals4Moms have the potential to transform how nutrition is integrated into perinatal care.

## Data Availability

All data produced in the present study are available upon reasonable request to the authors.

## Human Rights

All procedures performed in studies involving human participants were in accordance with the ethical standards of the institutional research committee and with the 1964 Helsinki declaration and its later amendments or comparable ethical standards. The study was approved by the UConn Health Institutional Review Board.

## Informed Consent

Informed consent was obtained from all individual participants in the study.

## Clinical Trials Registration #

NCT06227247

## Welfare of Animals

n/a

## Transparency statements

The study was pre-registered at https://clinicaltrials.gov (NCT06227247). Survey instruments and interview guides may be available by emailing Dr. Molly Waring at.

## Acknowledgements

We would like to thank the clinical staff who supported recruitment and the graduate and undergraduate students who assisted with data collection and data management.

## Online Supplemental Appendix

**Supplemental Table 1:** Additional characteristics of pregnant women with gestational diabetes (GDM), overall and by study condition, n (%) or M±SD.

|  | Overall<br>(n=8) | Meals4Moms<br>(n=4) | Usual care<br>(n=4) |
| --- | --- | --- | --- |
| Marital status |  |  |  |
| Married | 7 (88) | 3 (75) | 4 (100) |
| Living with partner | 1 (13) | 1 (25) | - |
| Number of other adults in household |  |  |  |
| 1 other adult | 7 (88) | 3 (75) | 4 (100) |
| 2 other adults | 1 (13) | 1 (25) | - |
| Pre-pregnancy BMI |  |  |  |
| Underweight | - | - | - |
| Normal weight | 3 (38) | 1 (25) | 2 (50) |
| Overweight | 2 (25) | 1 (25) | 1 (25) |
| Obesity | 3 (38) | 2 (50) | 1 (25) |
| Perceived diet quality |  |  |  |
| Poor | - | - | - |
| Fair | 1 (13) | 1 (25) | - |
| Good | 4 (50) | 1 (25) | 3 (75) |
| Very good | 3 (38) | 2 (50) | 1 (25) |
| Excellent | - | - | - |
| Ever used meal kit delivery service | 4 (50) | 3 (75) | 1 (25) |
| Used meal kit delivery service in this pregnancy ** | 1 (25) | - | 1 (100) |
| Has a smartphone | 8 (100) | 4 (100) | 4 (100) |
| Ever used Wi-Fi scale | 1 (13) | - | 1 (25) |
| Ever used a fitness tracker to track steps or exercise (e.g., Fitbit) | 7 (88) | 3 (75) | 4 (100) |
| Currently subscribes to podcasts | 3 (38) | 2 (50) | 1 (25) |
| How often listened to podcast in past 7 days ** |  |  |  |
| Never | - | - | - |
| Once | 1 (33) | 1 (50) | - |
| More than once but not every day | 2 (67) | 1 (50) | 1 (100) |
| Every day | - | - | - |

\* Missing data: n=1 for any vigorous physical activity in past 7 days at baseline
\*\* Among users of this technology or service

**Supplemental Table 2:**
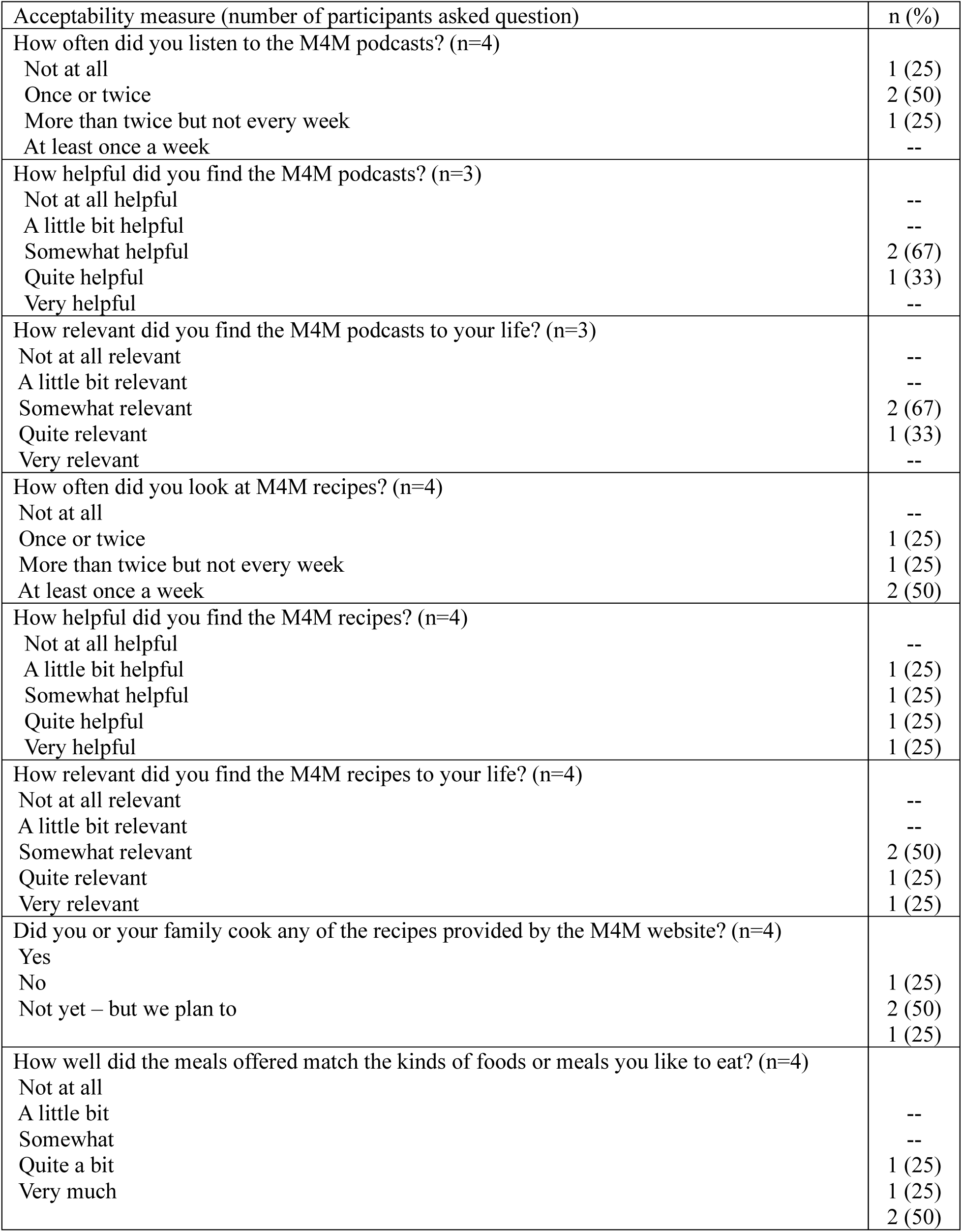
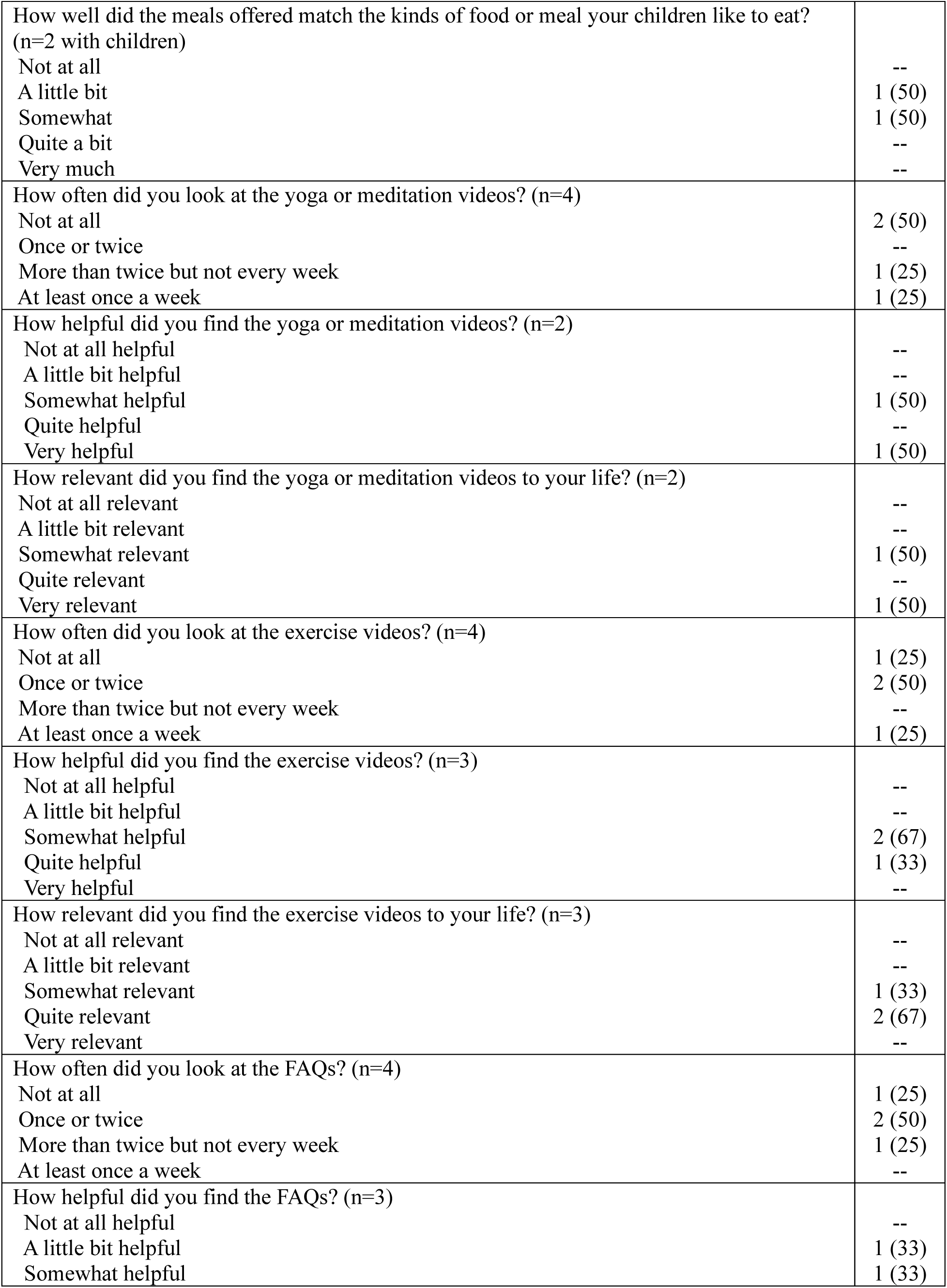
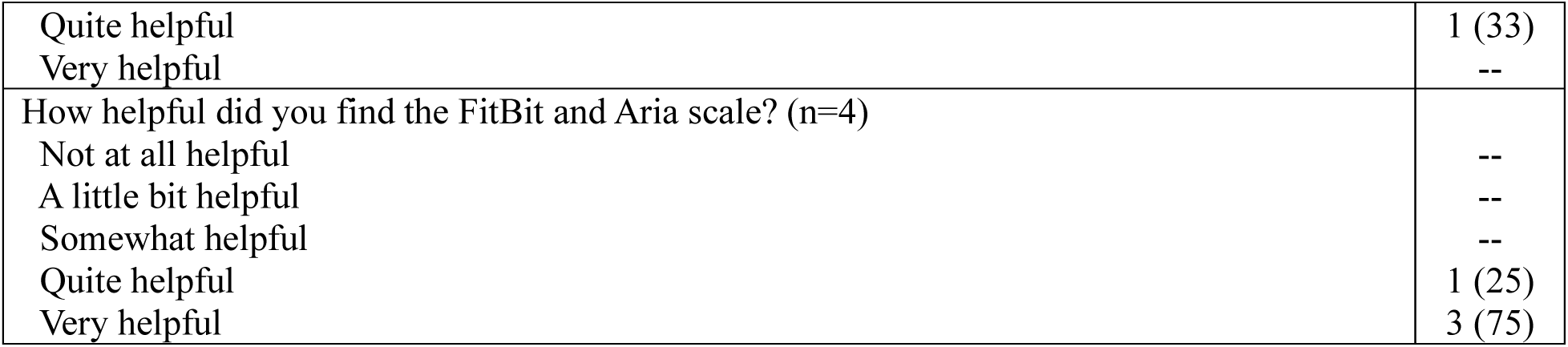
Measures of acceptability of the Meals4Moms program.

